# Cerebral hemodynamics response to dual-task paradigms in older adults: A systematic review and meta-analysis

**DOI:** 10.1101/2022.11.08.22282071

**Authors:** Kulvara Lapanan, Phunsuk Kantha, Gallayaporn Nantachai, Solaphat Hemrungrojn, Michael Maes

**Affiliations:** Department of Psychiatry, Faculty of Medicine, Chulalongkorn University, Bangkok, Thailand; Cognitive Fitness and Biopsychiatry Technology Research Unit, Faculty of Medicine, Chulalongkorn University, Bangkok, Thailand; School and Graduate Institute of Physical Therapy, College of Medicine, National Taiwan University, Taipei, Taiwan; Somdet Phra Sungharaj Nyanasumvara Geriatric Hospital, Department of Medical Services, Ministry of Public Health, Chon Buri Province, Thailand; Cognitive Impairment and Dementia Research Unit, Department of Psychiatry, Faculty of Medicine, Chulalongkorn University, Bangkok, Thailand; Deakin University, IMPACT - the Institute for Mental and Physical Health and Clinical Translation, School of Medicine, Barwon Health, Geelong, Australia; Department of Psychiatry, Medical University of Plovdiv, Plovdiv, Bulgaria

**Keywords:** Dual-task, Cognition, fNIRS, Executive function, Cerebral hemodynamics, Older adults

## Abstract

**Background:** Functional near-infrared spectroscopy (fNIRS) is a method to measure cerebral hemodynamics response. Changes in the prefrontal cortex during dual-tasking help to identify the risk of falling, particularly in older adults.

**Aims:** To systematically review and meta-analyze the effects of dual-task paradigms on cerebral hemodynamics in older adults.

**Methods:** The search was conducted in PubMed, Scopus, and Web of Science. A total of 22 studies comprising 1,841 older adults were included in the meta-analysis.

**Results:** Inhibitory control and working memory tasks significantly increased HbO_2_ in the PFC by 0.54 (*p* < 0.01, 95%CI = 0.36 – 0.72) and 0.13 (*p* < 0.01, 95%CI = 0.08 – 0.18) μmol/L, respectively. Overall, HbO_2_ was significantly increased during dual-task paradigms by 0.36 μmol/L (*P* < 0.01, 95%CI = 0.27 – 0.45). However, dual-task paradigms did not change HbR in the PFC (*P* = 0.14, 95%CI = -0.05 – 0.01).

**Conclusion:** Cognitive tasks related to inhibitory control required greater cognitive demands, indicating higher PFC activation during dual-task walking in older adults. This finding emphasizes the significance of assessing hemodynamic responses under dual-task paradigms to detect the risk of falling in older adults at an early stage.

## 1. Introduction

Aging causes deterioration in brain structure and function, which leads to a gradual decline in cognitive function (Li et al., 2001, Lindenberger et al., 2000, Fraser et al., 2017, Holtzer et al., 2016, Grady, 2000, Holtzer et al., 2011). Therefore, older adults show a decline in cognitive performance relative to younger adults (Li et al., 2001, Lindenberger et al., 2000, Holtzer et al., 2011). The significance of cognitive performance in older persons, particularly in terms of walking performance, is well-established (Verghese et al., 2002). Walking is a motor task that has been suggested to be an automatic movement influenced by cognitive function and motor control mechanisms (Abbott et al., 2004, Atkinson et al., 2007, Verghese et al., 2002, Verghese et al., 2007, Weuve et al., 2004). Higher cognitive demands (e.g., simultaneous walking and talking) are associated with slower walking speeds (Holtzer et al., 2016, Beauchet et al., 2008, Bootsma-van der Wiel et al., 2003). In addition, the difficulty to maintain a conversation while walking (i.e., stopping walking when talking) is predictive of future falls in older persons.(Beauchet et al., 2009, de Hoon et al., 2003, Young et al., 2016). This suggests that cognitive function plays an important role when performing motor tasks in conjunction with cognitive tasks, especially in older adults.

Dual-task is defined as the simultaneous execution of two distinct tasks (Lindenberger et al., 2000). For example, in daily life, it is common to encounter situations in which a motor task (e.g., standing or walking) is performed along with a cognitive task (e.g., thinking, or talking). Reduced ability to perform dual-task is the leading cause of postural instability and increases the risk of falling in older adults (Hall et al., 2010, Hausdorff, 2005). In older adults, falls are directly associated with bone fractures, resulting in substantial complications such as increased risk of disability, lower quality of life, and higher mortality (Kannus et al., 2005). Furthermore, older adults are more likely to fall when undertaking dual-tasking while walking compared to walking only (Lundin-Olsson et al., 1997). Therefore, understanding the mechanism that underpin dual-task paradigms in older adults is crucial.

The prefrontal cortex (PFC) is the cerebral cortex located in the front part of the brain, which is primarily responsible for executive function, including inhibitory control, working memory, and cognitive flexibility (Diamond, 2013). The executive function is associated with processing and selecting attention for responding when performing cognitive tasks (Rosso et al., 2017, Verghese et al., 2002). Individuals who have higher PFC activity frequently exert more effort to complete tasks, indicating that they have less cognitive function (Quaresima and Ferrari, 2019). Thus, in terms of brain activity, the PFC is the most common brain area to measure executive function when performing cognitive tasks.

Functional Near-Infrared Spectroscopy (fNIRS) is a non-invasive brain imaging method that uses infrared light to measure cerebral hemodynamics (Karim et al., 2013). fNIRS takes advantage of the brain absorption of infrared light, which is different between the spectra of oxygenated hemoglobin (HbO_2_) and deoxygenated hemoglobin (HbR) (Quaresima and Ferrari, 2019). The fluctuations in HbO_2_ and HbR have been suggested for use as a measure of cortical activity (Quaresima and Ferrari, 2019). For instance, the increased demands on the brain usually cause an influx of HbO_2_ and an efflux of HbR (Quaresima and Ferrari, 2019). Moreover, because of its portability and no restriction on motion, fNIRS is a suitable technique for determining cortical activity during any physical activity (Pinti et al., 2020).

Recently, fNIRS has been extensively used to investigate the cerebral hemodynamics of PFC in older adults (Beurskens et al., 2014, Chen et al., 2017, Corp et al., 2018, Hassan et al., 2020, Hawkins et al., 2018, Hernandez et al., 2020, Hoang et al., 2022, Holtzer and Izzetoglu, 2020, Holtzer et al., 2011, Holtzer et al., 2020, Holtzer et al., 2022, Maidan et al., 2016, Marusic et al., 2019, Ross et al., 2021, Rosso et al., 2017, Salzman et al., 2021b, Teo et al., 2021). Studies have revealed that PFC activity may increase, decrease, or show no change when performing dual-tasks (Beurskens et al., 2014, Holtzer et al., 2022, Izzetoglu and Holtzer, 2020, Maidan et al., 2016, Mirelman et al., 2017, Salzman et al., 2021b, Stuart et al., 2019). Because of these discrepancies, there is no consensus on how dual-tasks may affect cerebral hemodynamics of the PFC in fNIRS studies. Therefore, this study aimed to systematically determine the effect of dual-task paradigms on the change in the cerebral hemodynamics (including HbO_2_ and HbR) in older adults. Researchers and professionals might benefit from understanding how cerebral hemodynamics changes when performing dual-tasks since this information might be crucial for explaining falls in the elderly.

### 2. Materials and Methods

This study was conducted according to the guidelines in the Preferred Reporting Items for Systematic Reviews and Meta-analysis (PRISMA) statement (Page et al., 2021).

### 2.1 Data sources and search strategy

The search was conducted in the PubMed, Scopus, and Web of Science databases from inception until September 2022. A search strategy was implemented to find the study that investigated the effect of dual-task on cerebral hemodynamics in older adults. The search strategy for all databases is shown in the supplementary materials.

### 2.2 Eligibility criteria

The inclusion criteria for this study were as follows: (a) population: older adults (age above 50 years old) (Sabharwal et al., 2015); (b) intervention: any cognitive task (i.e., dual-task); (c) comparison: any motor task (i.e., single-task); (d) outcome: cerebral hemodynamics. We excluded studies in which the population had cognitive impairment (e.g., dementia and Alzheimer’s) and motor deficits (e.g., stroke). Intervention studies with more than two task integrations were excluded. We also excluded non-English language or non-original research studies.

### 2.3 Study Selection

All included studies were imported into Endnote 20 (Clarivate Analytics, Boston, USA) to remove the duplicates. After the removal of duplicate studies, two independent reviewers (KL and PK) reviewed the remaining studies in accordance with the eligibility criteria on the titles and abstracts. Disagreements between the two independent reviewers (KL and PK) about study selection were resolved by discussion with the third independent reviewer (GN).

### 2.4 Data Extraction

Data from all included studies were extracted and summarized in the table (see **Table 1**) by two independent reviewers (KL and PK). Two independent reviewers (KL and PK) retrieved the following data from each included study: (a) first author and year of publication; (b) sample size and population information; (c) single-task description; (d) dual-task description; and (e) all outcome variables. In case the data were unclear or incomplete, we contacted the first or corresponding author of the study by email. If we did not receive a response by email, the data was estimated by using a program custom-written in the MATLAB R2020a software (MathWorks, Natrick, MA, USA) (Shi et al., 2020, Luo et al., 2018). When the two independent reviewers (KL and PK) disagreed on data extraction, the third independent reviewer (GN) was invited to a meeting to find an agreement on adjudication.

**Table 1.**
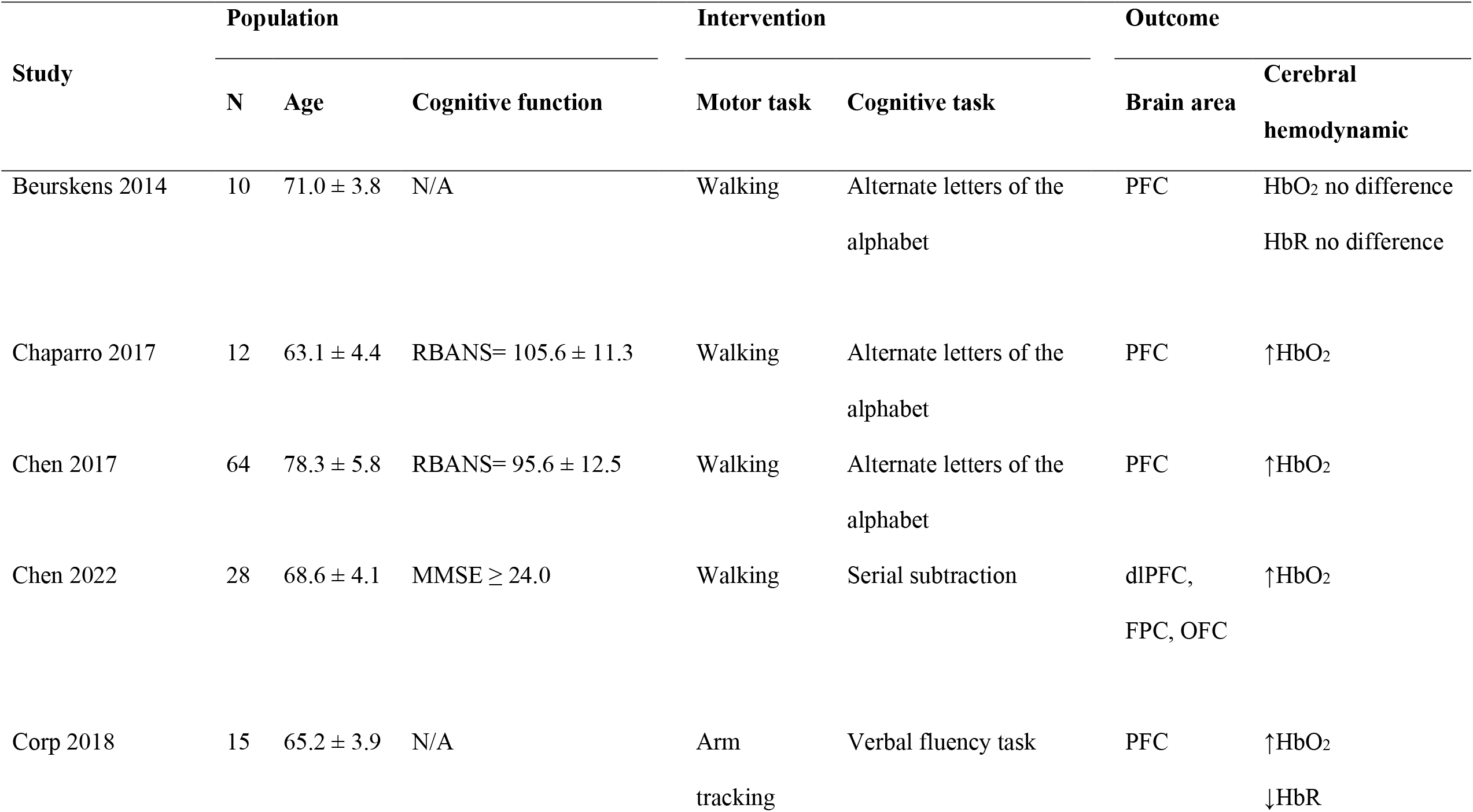
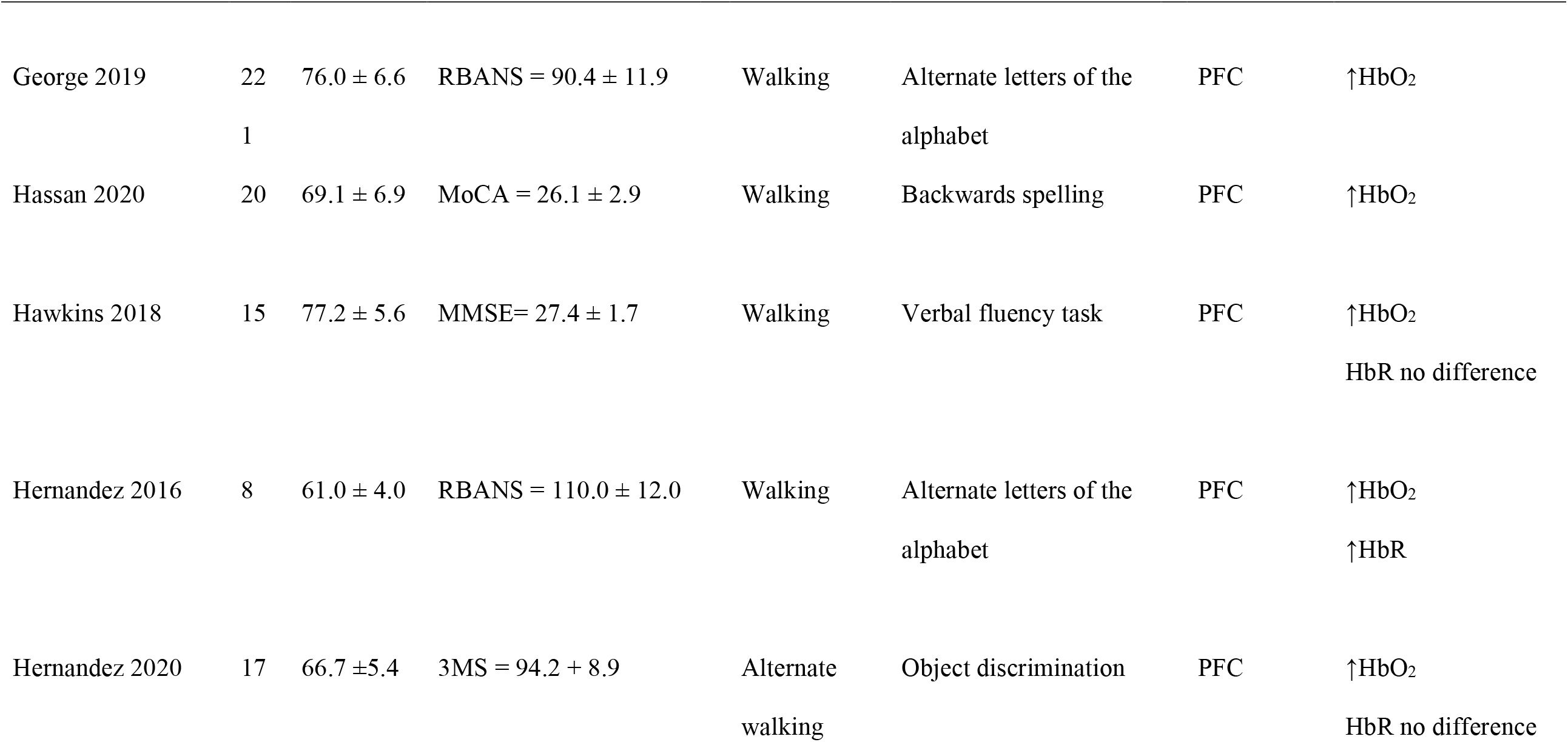
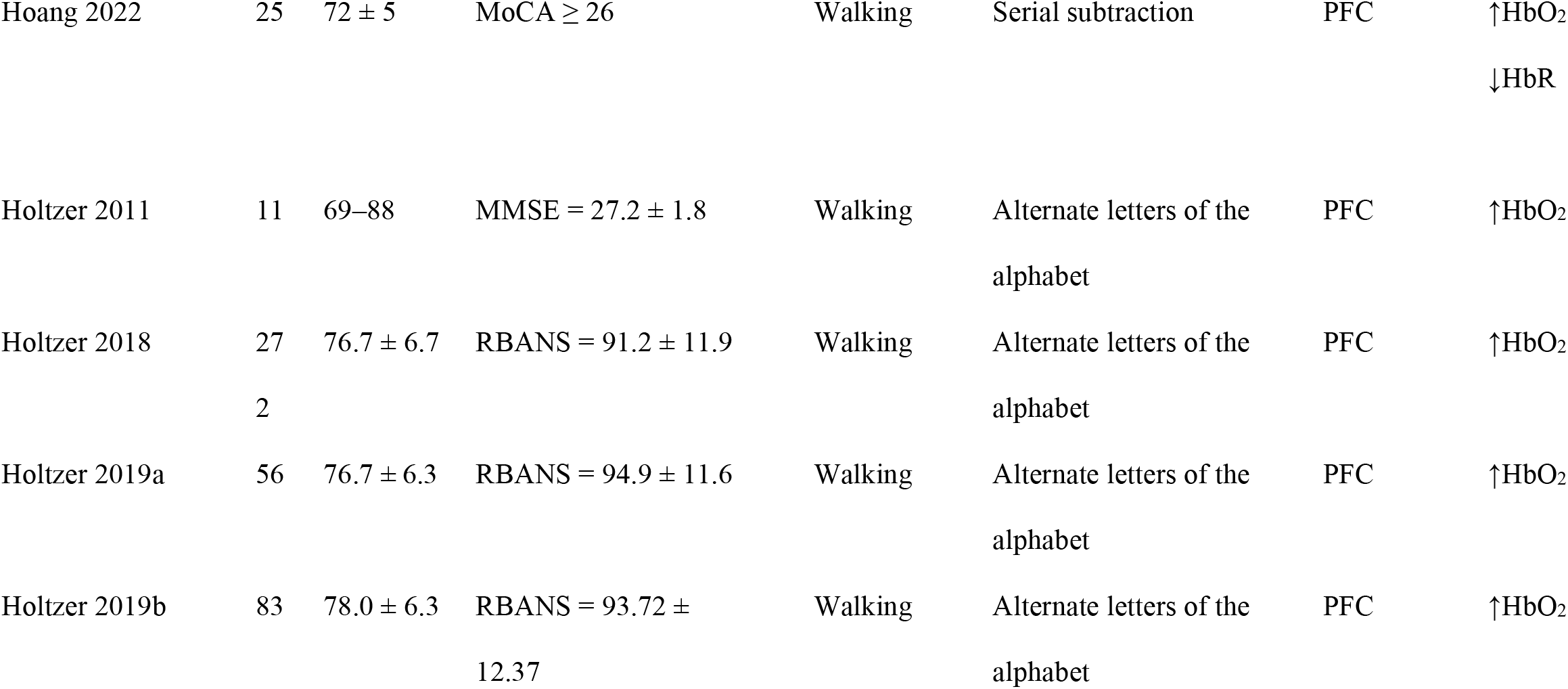
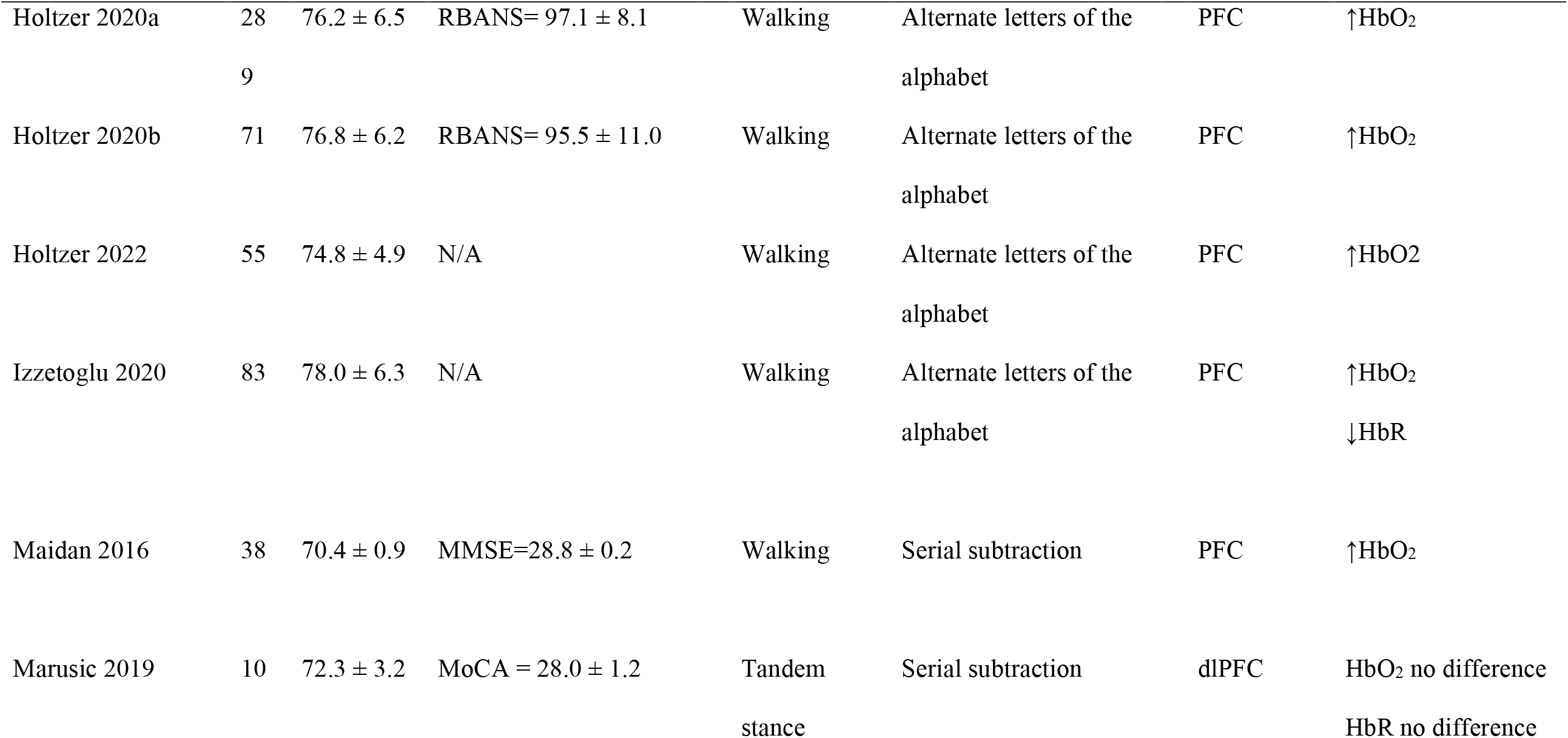
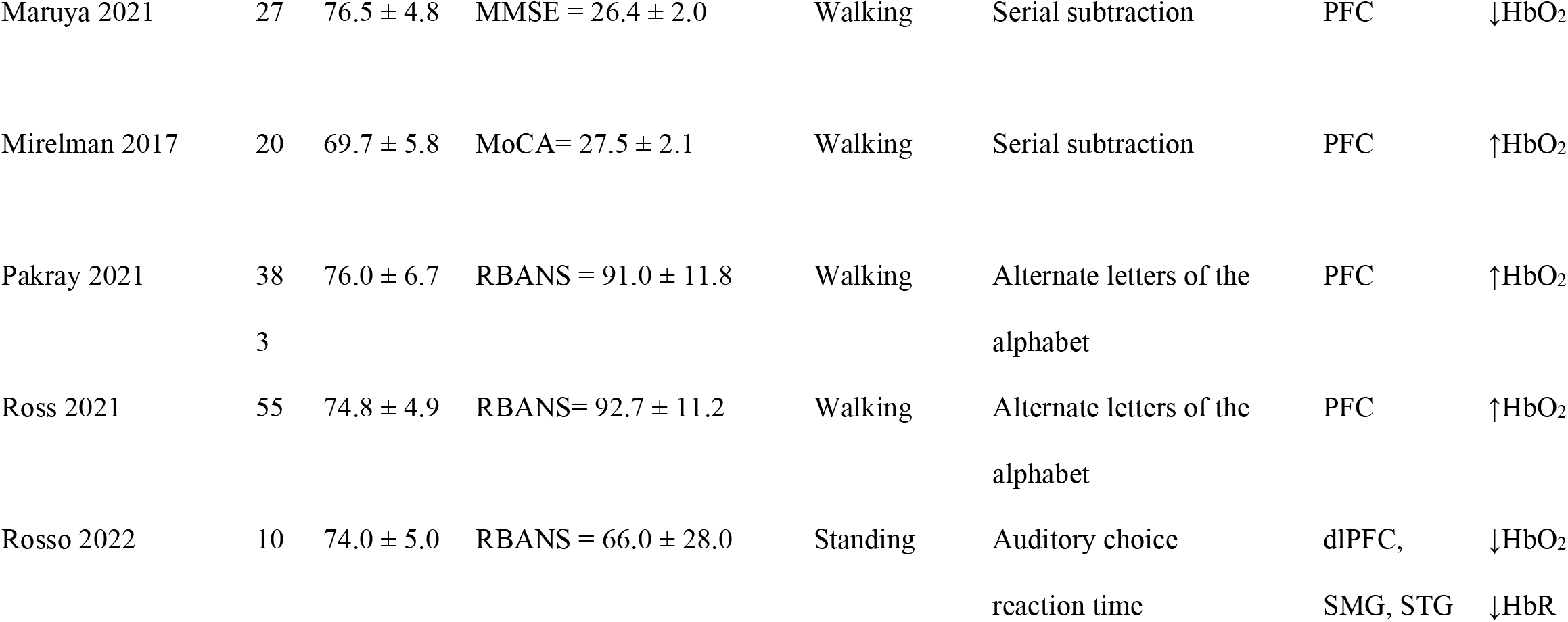
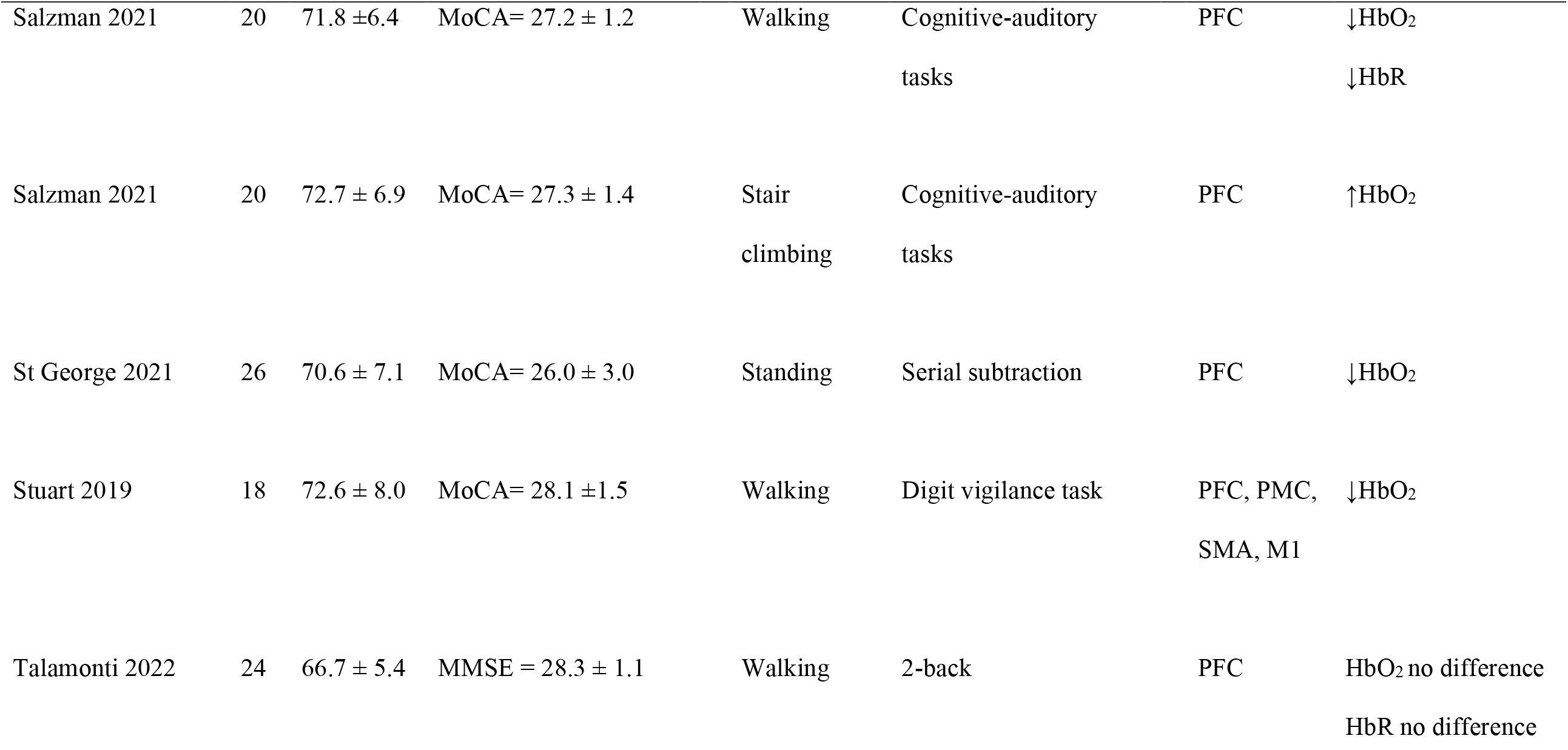
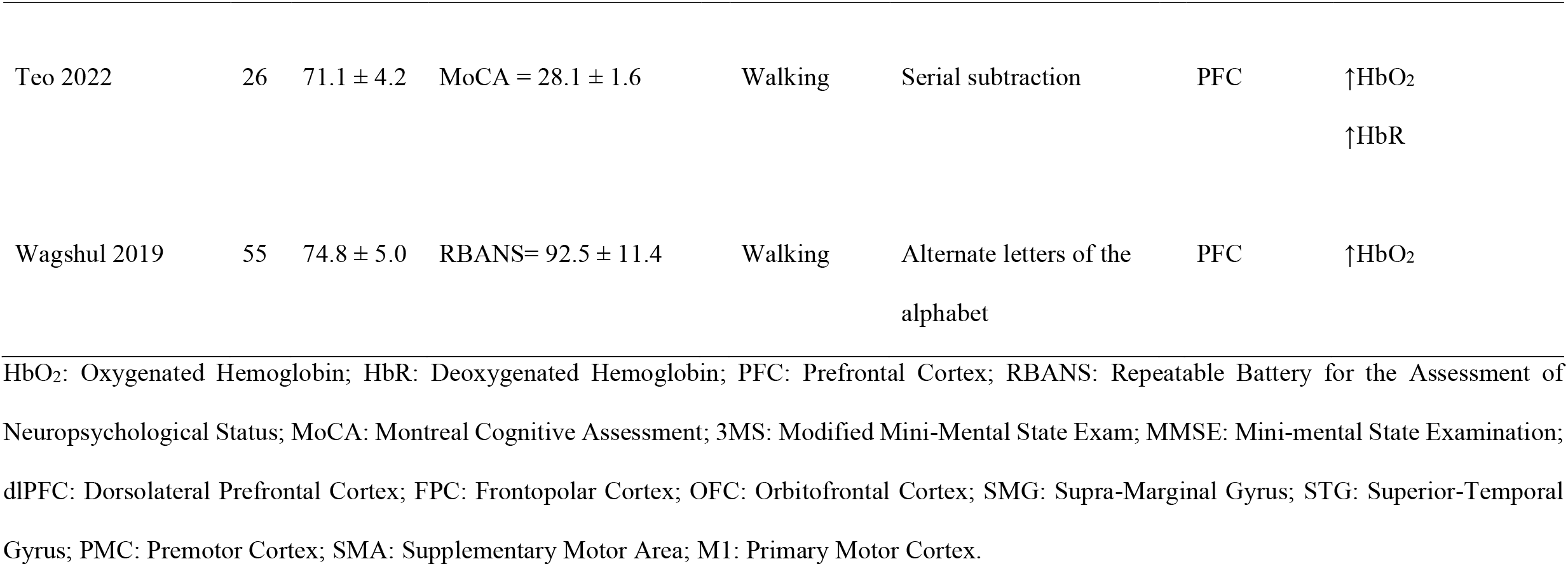
Summary of the included study characteristics.

### 2.5 Risk of bias assessment

Two independent reviewers (KL and PK) assessed the risk of bias of the included studies according to the Risk of Bias In Non-Randomized Studies-of Interventions (ROBINS-I) (Sterne et al., 2016). ROBINS-I consists of the following domains: (a) bias due to confounding; (b) bias in selection of participants; (c) bias in classification of interventions; (d) bias due to deviations from intended interventions; (e) bias due to missing data; (f) bias in measurement of outcomes; and (g) bias in selection of the reported result. The judgments for each domain were low risk of bias, unclear risk of bias, and high risk of bias. Disagreements between the two independent reviewers (KL and PK) about the risk of bias were also resolved by discussion with the third independent reviewer (GN).

### 2.6 Synthesis of results

The Review Manager v.5.3 (RevMan, The Nordic Cochrane Centre, Cochrane Collaboration, Copenhagen, Denmark) was used to determine the cerebral hemodynamics between single-task and dual-task. Data synthesis was conducted when the included studies reported a similar primary outcome in mean ± SD for at least 3 studies (Page et al., 2021). Subgroup analysis was conducted to compare the effects of different types of cognitive tasks. The cognitive tasks were categorized in accordance with their respective executive functions (Diamond, 2013): a) inhibitory control tasks: the tasks that suppress or countermand before response; b) working memory tasks: the tasks that provide the starting information and require the information to be manipulated based on conditions; and c) cognitive flexibility tasks: the tasks that allow to choose between two (or more than two) different responses. This meta-analysis used weighted mean differences (WMD) with 95% confidence intervals (95% CI) because the outcome was presented in the same units (Page et al., 2021). The I^2^ value was used to define the heterogeneity of the outcome (Page et al., 2021). I^2^ values of more than 50% were used to indicate heterogeneity. The alpha level was set at 0.05 for statistical significance.

## 3. Results

### 3.1 Search results

The PRISMA flowchart shows a summary of the study selection process, as illustrated in **Fig. 1**. The initial search identified a total of 1,421 studies after searching three databases, one of which was an additional study from a manual search (Hassan et al., 2020). 920 studies remained after the removal of duplicates. The remaining studies were then screened by title and abstract. After the screening, 73 studies were assessed by full-text reviews. We excluded a total of 40 studies for the following reasons: non-original research studies (n = 5), individuals with cognitive impairment (n = 5), interventions involving more than two tasks (n = 24), and not comparing with single-task (n = 6). Finally, 33 eligible studies (Beurskens et al., 2014, Chaparro et al., 2017, Chen et al., 2017, Chen et al., 2022, Corp et al., 2018, George et al., 2019, Hassan et al., 2020, Hawkins et al., 2018, Hernandez et al., 2020, Hernandez et al., 2016, Hoang et al., 2022, Holtzer et al., 2018, Holtzer and Izzetoglu, 2020, Holtzer et al., 2019a, Holtzer et al., 2019b, Holtzer et al., 2011, Holtzer et al., 2020, Holtzer et al., 2022, Izzetoglu and Holtzer, 2020, Maidan et al., 2016, Marusic et al., 2019, Mirelman et al., 2017, Pakray et al., 2021, Ross et al., 2021, Rosso et al., 2017, Sabharwal et al., 2015, Salzman et al., 2021a, Salzman et al., 2021b, St George et al., 2021, Stuart et al., 2019, Talamonti et al., 2022, Teo et al., 2021, Wagshul et al., 2019, Maruya et al., 2021) were included in the systematic review. However, because of limitations in data extraction, 22 studies (Beurskens et al., 2014, Chen et al., 2022, Corp et al., 2018, Hernandez et al., 2020, Marusic et al., 2019, Pakray et al., 2021, Rosso et al., 2017, Salzman et al., 2021a, Salzman et al., 2021b, St George et al., 2021, Stuart et al., 2019, Talamonti et al., 2022) were included in the quantitative synthesis.

**Fig. 1.**
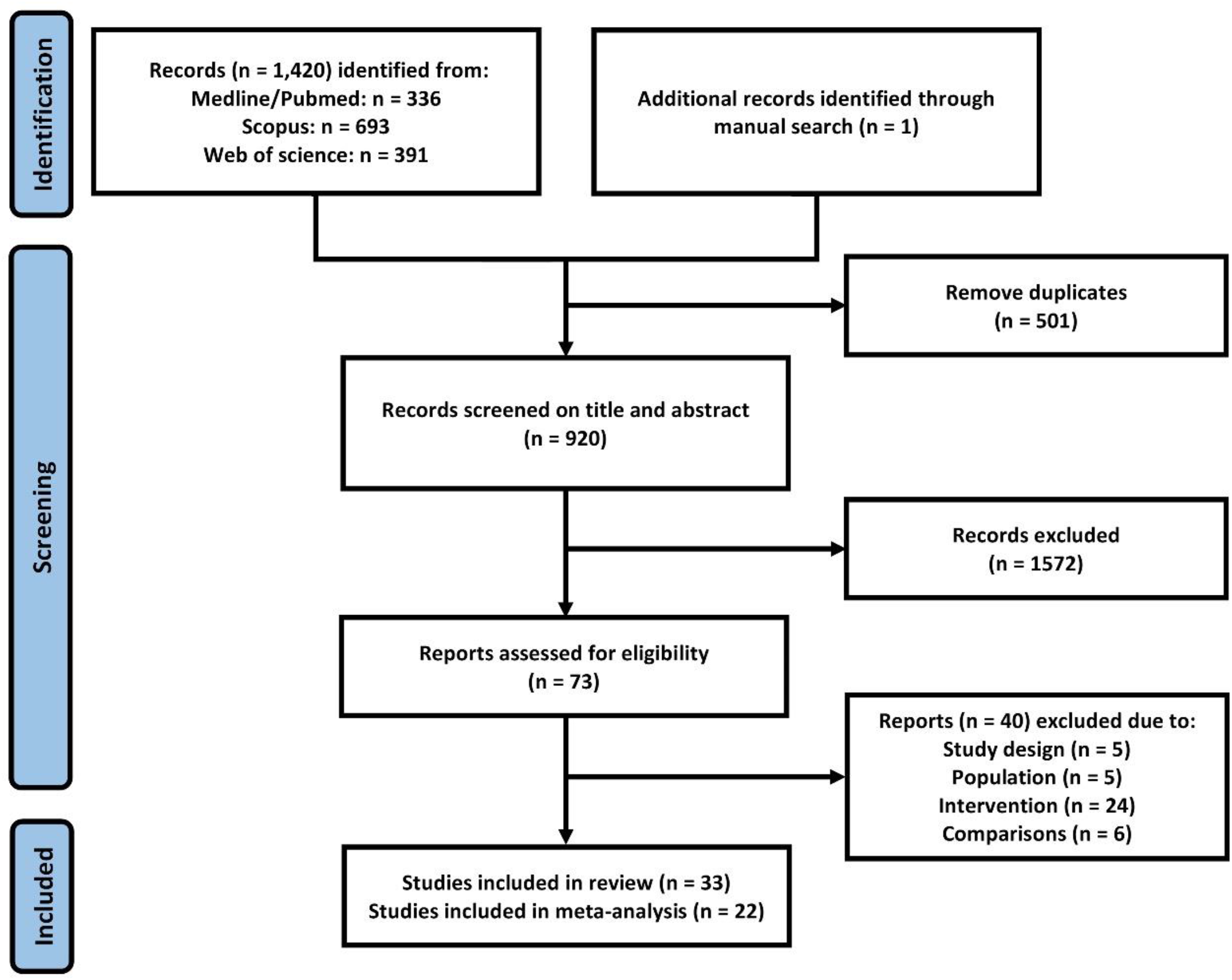
PRISMA flow diagram summarizing study screening and selection for review.

### 3.2 Study characteristics

The included studies were published between 2011 and 2022. The characteristics of the included studies are summarized in **Table 1**. A total of 2,087 older adults were recruited from 33 studies (Beurskens et al., 2014, Chaparro et al., 2017, Chen et al., 2017, Chen et al., 2022, Corp et al., 2018, George et al., 2019, Hassan et al., 2020, Hawkins et al., 2018, Hernandez et al., 2020, Hernandez et al., 2016, Hoang et al., 2022, Holtzer et al., 2018, Holtzer and Izzetoglu, 2020, Holtzer et al., 2019a, Holtzer et al., 2019b, Holtzer et al., 2011, Holtzer et al., 2020, Holtzer et al., 2022, Izzetoglu and Holtzer, 2020, Maidan et al., 2016, Marusic et al., 2019, Mirelman et al., 2017, Pakray et al., 2021, Ross et al., 2021, Rosso et al., 2017, Sabharwal et al., 2015, Salzman et al., 2021a, Salzman et al., 2021b, St George et al., 2021, Stuart et al., 2019, Talamonti et al., 2022, Teo et al., 2021, Wagshul et al., 2019, Maruya et al., 2021), with 1,841 older adults from 22 studies of them included in the meta-analysis. The age of participants ranged from 61 to 78 years old (mean 75.26 ± 6.11 years). Most of the included studies clearly stated that the older adults had no cognitive impairment, except for 4 studies (Beurskens et al., 2014, Holtzer et al., 2022, Izzetoglu and Holtzer, 2020, Corp et al., 2018) that did not report. For single-task paradigm, all of the included studies used a motor task including walking in 28 studies (Beurskens et al., 2014, Chaparro et al., 2017, Chen et al., 2017, Chen et al., 2022, George et al., 2019, Hassan et al., 2020, Hawkins et al., 2018, Hernandez et al., 2020, Hernandez et al., 2016, Hoang et al., 2022, Holtzer et al., 2018, Holtzer and Izzetoglu, 2020, Holtzer et al., 2019a, Holtzer et al., 2019b, Holtzer et al., 2011, Holtzer et al., 2020, Holtzer et al., 2022, Izzetoglu and Holtzer, 2020, Maidan et al., 2016, Maruya et al., 2021, Mirelman et al., 2017, Pakray et al., 2021, Ross et al., 2021, Salzman et al., 2021b, Stuart et al., 2019, Talamonti et al., 2022, Teo et al., 2021, Wagshul et al., 2019), standing in 3 studies (Marusic et al., 2019, St George et al., 2021, Rosso et al., 2017), arm tracking in 1 study (Corp et al., 2018), and stair climbing in 1 study (Salzman et al., 2021a). For dual-task paradigm, cognitive tasks were performed as follows: 19 studies (Chaparro et al., 2017, Chen et al., 2017, George et al., 2019, Hernandez et al., 2016, Hoang et al., 2022, Holtzer et al., 2018, Holtzer and Izzetoglu, 2020, Holtzer et al., 2019a, Holtzer et al., 2019b, Holtzer et al., 2011, Holtzer et al., 2020, Holtzer et al., 2022, Izzetoglu and Holtzer, 2020, Ross et al., 2021, Wagshul et al., 2019, Rosso et al., 2017, Salzman et al., 2021a, Salzman et al., 2021b, Beurskens et al., 2014, Corp et al., 2018, Hernandez et al., 2020, Marusic et al., 2019, Pakray et al., 2021, Stuart et al., 2019, Talamonti et al., 2022, Chen et al., 2022, Hassan et al., 2020, Hawkins et al., 2018) used inhibitory control tasks (i.e., alternate letters of alphabet and auditory choice reaction), 14 studies (Hassan et al., 2020, Hawkins et al., 2018, Hoang et al., 2022, Maidan et al., 2016, Maruya et al., 2021, Mirelman et al., 2017, Teo et al., 2021, Talamonti et al., 2022, Corp et al., 2018, Beurskens et al., 2014, Hernandez et al., 2020, Marusic et al., 2019, Pakray et al., 2021, Stuart et al., 2019, Chen et al., 2022, St George et al., 2021) used working memory tasks (i.e., backward spelling, serial subtraction, n-back and verbal fluency task). None of the included studies used cognitive flexibility tasks as cognitive task. All of the included studies mainly investigated the changes in cerebral hemodynamics in the PFC.

### 3.3 The effect of dual-task paradigms on oxygenated hemoglobin (HbO_2_)

All of the data synthesis used walking as a single-task (100%), while the dual-task paradigms were inhibitory control tasks (77%) and working memory (23%). For subgroup analysis, in inhibitory control task, the majority of studies favor dual-task paradigm, but 1 study (Maruya et al., 2021) favor single-task paradigm. A total of 15 studies (Chaparro et al., 2017, Chen et al., 2022, George et al., 2019, Hernandez et al., 2016, Holtzer et al., 2018, Holtzer and Izzetoglu, 2020, Holtzer et al., 2019a, Holtzer et al., 2019b, Holtzer et al., 2011, Holtzer et al., 2020, Holtzer et al., 2022, Izzetoglu and Holtzer, 2020, Ross et al., 2021, Salzman et al., 2021b, Wagshul et al., 2019, Chen et al., 2017) involving 1,355 older adults were analyzed in order to determine the effect of inhibitory control task on HbO_2_. The data synthesis revealed that inhibitory control tasks significantly increased HbO_2_ concentration in the PFC by 0.54 μmol/L (Z = 5.85, *p* < 0.01, 95% CI = 0.36 to 0.72, I^2^ = 99%). Meanwhile, in working memory task, a total of 7 studies (Hassan et al., 2020, Hawkins et al., 2018, Hoang et al., 2022, Maidan et al., 2016, Maruya et al., 2021, Mirelman et al., 2017, Teo et al., 2021) involving 171 older adults were analyzed in order to determine the effect of working memory task on HbO_2_. The data synthesis revealed that working memory tasks significantly increased HbO_2_ concentrations in the PFC by 0.13 μmol/L (Z = 5.05, *p* < 0.01, 95% CI = 0.08 to 0.18, I^2^ = 89%). Therefore, a total of 22 studies (Hassan et al., 2020, Hawkins et al., 2018, Hoang et al., 2022, Maidan et al., 2016, Maruya et al., 2021, Mirelman et al., 2017, Teo et al., 2021, Chaparro et al., 2017, Chen et al., 2022, George et al., 2019, Hernandez et al., 2016, Holtzer et al., 2018, Holtzer and Izzetoglu, 2020, Holtzer et al., 2019a, Holtzer et al., 2019b, Holtzer et al., 2011, Holtzer et al., 2020, Holtzer et al., 2022, Izzetoglu and Holtzer, 2020, Ross et al., 2021, Salzman et al., 2021b, Wagshul et al., 2019) involving 1,526 older adults were analyzed in order to determine the overall effect of dual-task paradigms on HbO_2_. Meta-analysis revealed that dual-task paradigms significantly increased HbO_2_ concentration in the PFC by 0.36 μmol/L (Z = 7.91, P < 0.01, 95% CI = 0.27 to 0.45, I^2^ = 99%). **Fig 2A** shows a forest plot displaying the effects of the dual-task paradigms on HbO_2_.

**Fig. 2.**
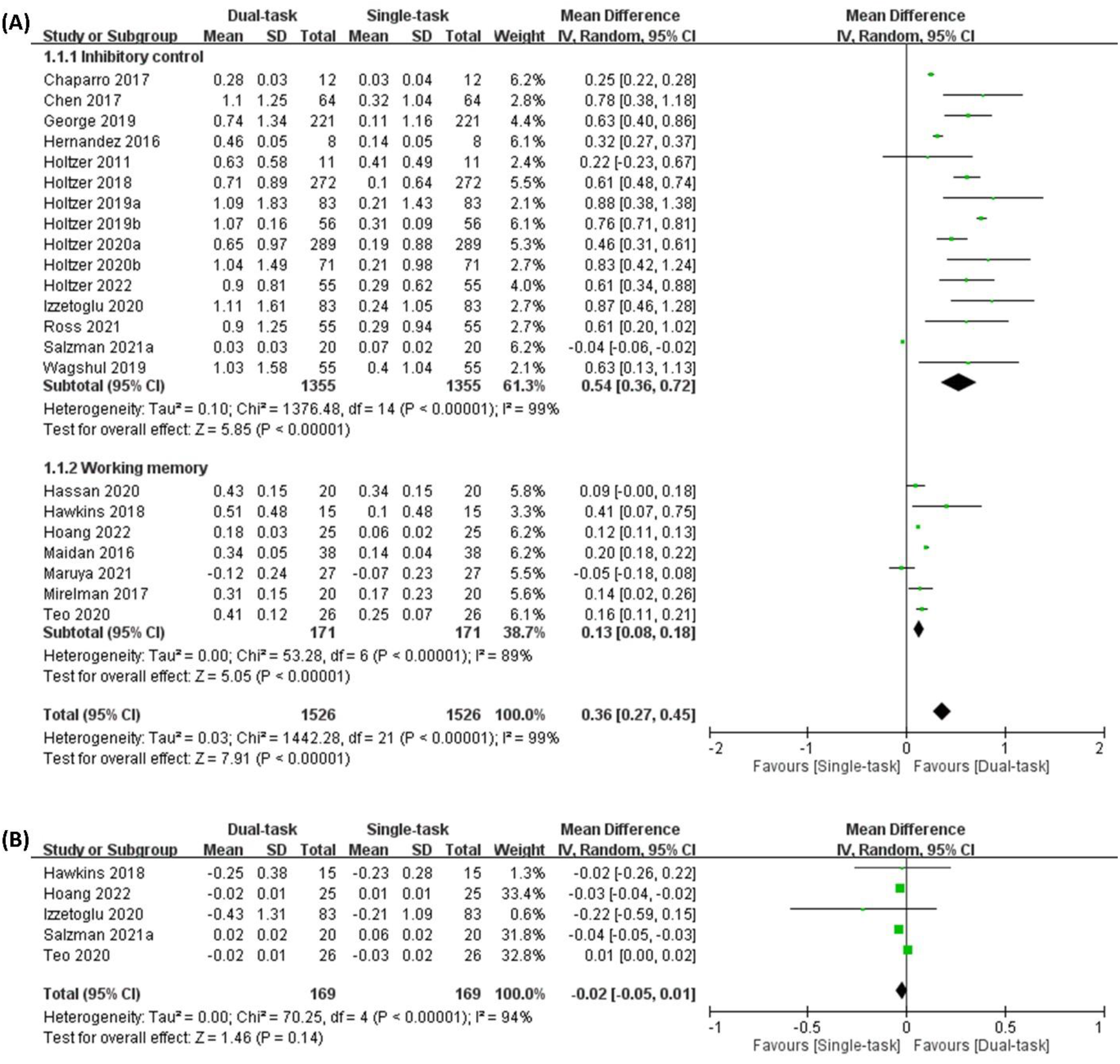
Forest plots showing weighted mean differences with 95% CI in the influence of dual-task paradigms on: **(A)** Oxygenated Hemoglobin and **(B)** Deoxygenated Hemoglobin.

### 3.4 The effect of dual-task paradigms on deoxygenated hemoglobin (HbR)

Due to the low number of included studies, the subgroup analysis was not conducted for the effect of dual-task paradigms on HbR. Most of the included studies favor single-task paradigm (Hawkins et al., 2018, Hoang et al., 2022, Izzetoglu and Holtzer, 2020, Salzman et al., 2021b), but only 1 study (Teo et al., 2021) favors dual-task paradigm. A total of 5 studies (Hawkins et al., 2018, Hoang et al., 2022, Izzetoglu and Holtzer, 2020, Salzman et al., 2021b, Teo et al., 2021) involving 169 older adults revealed that the dual-task paradigms did not change HbR in the PFC (WMD = - 0.02, Z = 1.46, *P* = 0.14, 95% CI = -0.05 to 0.01, I^2^ = 94%). **Fig. 2B** shows the forest plot of the influence of dual-task paradigms on HbR.

### 3.5 Risk of bias

4 studies (Beurskens et al., 2014, Holtzer et al., 2019b, Holtzer et al., 2022, Izzetoglu and Holtzer, 2020) out of the 33 studies (12%) demonstrated an unclear risk in the domain of confounding. 5 studies (15%) showed a high risk of bias in the classification of interventions (Corp et al., 2018, Marusic et al., 2019, Rosso et al., 2017, Salzman et al., 2021a, St George et al., 2021). 2 studies (6%) showed a high risk of bias in selection of the reported results (Rosso et al., 2017, Teo et al., 2021). However, all the included studies had a low risk of bias in the remaining domains (i.e., selection of participants, deviations from intended interventions, missing data, and measurement of outcomes). The risk of bias for all included studies is summarized in **Fig. 3**. The detailed risk of bias for each study regarding the seven domains is illustrated in **Fig. 4**.

**Fig. 3.**
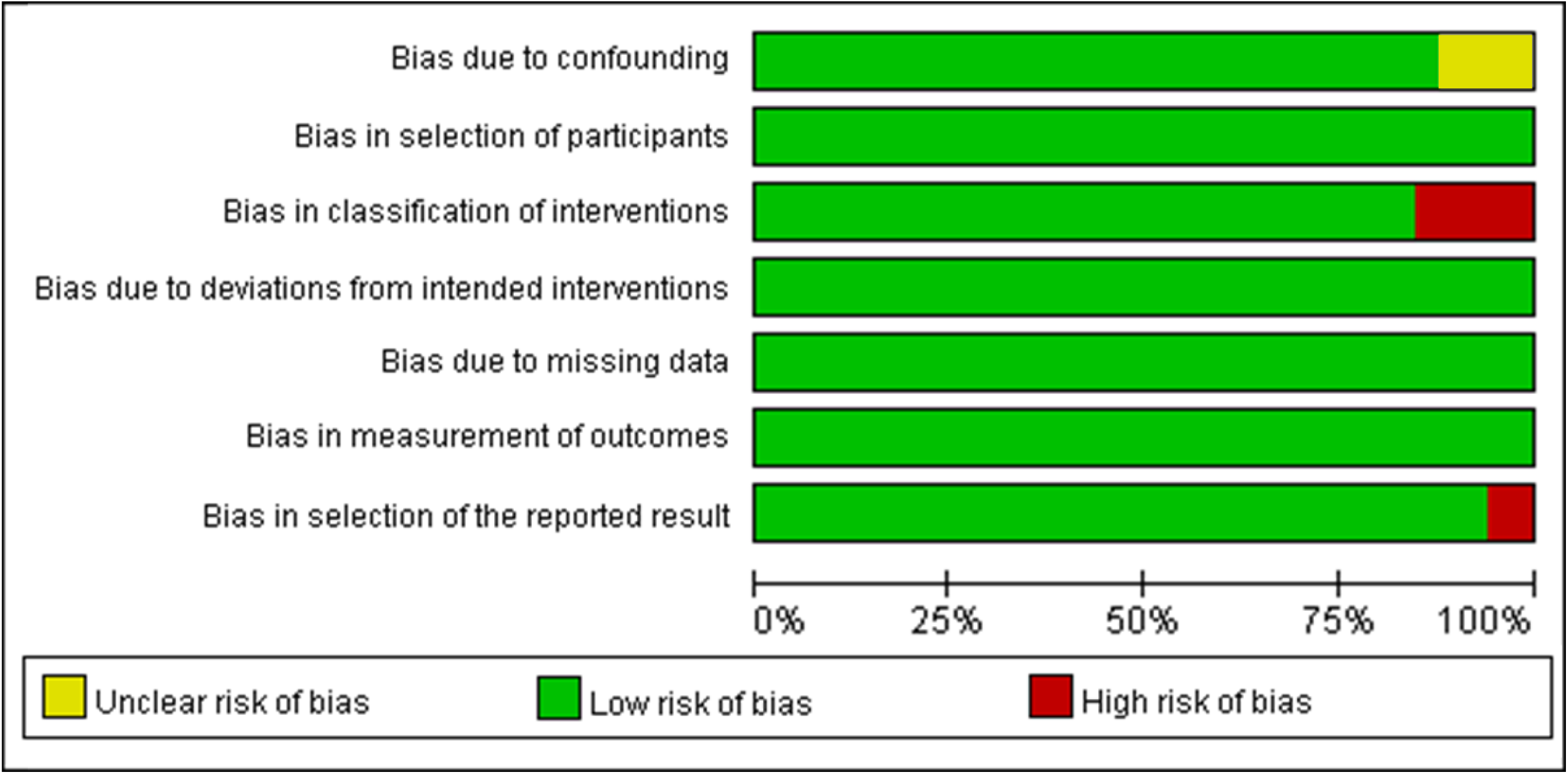
Summary risk of bias for all included studies.

**Fig. 4.**
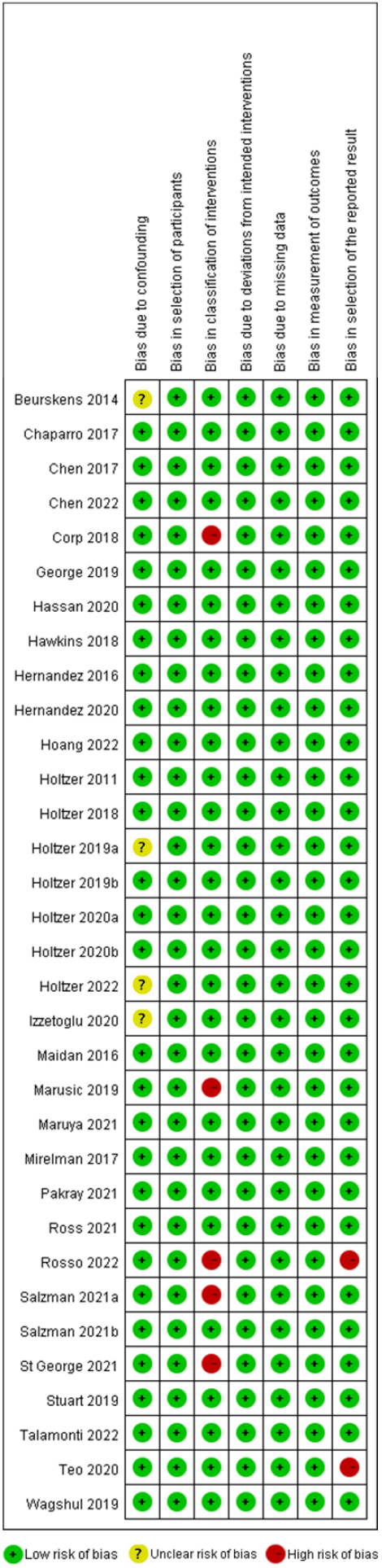
Detailed risk of bias for each study.

## 4. Discussion

This study is the first to systematically review and meta-analyze all research studies on changes in cerebral hemodynamics due to dual-task paradigms in older adults. Most of the included studies demonstrated a low risk of bias, indicating that only studies with good quality evidence were included in the data synthesis. According to the data synthesis, dual-task paradigms (both inhibitory control and working memory tasks) increase HbO_2_ concentration but have no effect on HbR in comparison to single-task in older adults. Specifically, the inhibitory control tasks were more likely to increase HbO_2_ than working memory tasks. Therefore, we suggested that cognitive tasks related to the inhibitory control tasks required greater cognitive demands, resulting in higher cortical activity during dual-task walking.

### 4.1 The effect of dual-task paradigms on oxygenated hemoglobin (HbO_2_)

Dual-task paradigms resulted in a higher HbO_2_ concentration in the PFC of older adults when compared with single-task paradigms. In the single-task, older adults also consume some oxygen when performing a motor task, such as walking alone. This was consistent with previous findings that the human brain utilizes oxygen for approximately 20% of the total oxygen consumption at rest (Talamonti et al., 2021). For multitasking, the brain requires substantially more oxygen to maintain optimal oxygen levels (i.e., dual-task paradigm) (Marusic et al., 2019, Maruya et al., 2021, Mihara et al., 2008). For this reason, dual-tasking necessitates adequate oxygenation for normal brain function (Marusic et al., 2019). In our data synthesis, older adults have higher HbO_2_ (i.e., higher oxygen consumption) in the bilateral PFC when performing dual-task, which suggests a greater reliance on executive functions. However, no prior study has delineated the precise amount of HbO_2_ increase that is associated with an increased risk of falling.

#### 4.1.1 Differential effects of cognitive tasks on PFC activation

The cognitive tasks in the included studies were used as a dual-task to increase cognitive demands during walking. The most common cognitive tasks were inhibitory control tasks (i.e., alternate letters of the alphabet and auditory choice reaction) and working memory tasks (i.e., serial subtraction, verbal fluency, and backwards spelling). Previous studies reported that the underlying mechanism of inhibitory control were associated with an increase in HbO_2_ concentration in the right PFC (Marusic et al., 2019, Hassan et al., 2020). Previous studies showed that the concentration of HbO_2_ in the left PFC increases when performing cognitive tasks requiring working memory (Cabeza and Nyberg, 2000, Gabrieli et al., 1998). Although the included studies did not independently report the HbO_2_ concentration in the left or right lobes of the PFC, we found that inhibitory control tasks have a greater effect on HbO_2_ as compared with working memory tasks. The possibility is that the cognitive tasks related to the right PFC activity require more cortical activity.

Most of the included studies showed an increase in HbO_2_ concentration during dual-task walking. However, some of the included studies showed a decrease in HbO_2_ when walking with cognitive tasks (i.e., auditory choice reaction time task (Rosso et al., 2017), visual task (Beurskens et al., 2014), alternating letters of alphabet (Izzetoglu and Holtzer, 2020) or cognitive-auditory task (Salzman et al., 2021b). The cognitive information processing hypothesis can explain the discrepancies (Atkinson et al., 2007). According to this theory, cognitive tasks might be recognized in the long-term memory, which has an advantageous effect in minimizing interference from dual-tasks (Callan et al., 2014). Hence, executive functions are constrained and reorganized in accordance with long-term memory. Moreover, a number of studies have demonstrated that the decrease in PFC activation is associated with automatic tasks (Dietrich and Audiffren, 2011, Wu et al., 2004). Nevertheless, this study does not provide a consensus regarding the decrease in HbO_2_ in older adults under dual-tasking.

### 4.2 The effect of dual-task paradigms on deoxygenated hemoglobin (HbR)

Our findings suggest that dual-task paradigms had no effect on HbR concentration in the PFC of older adults. Normally, a decrease in HbR concentration coincides with an increase in HbO_2_. As a result of previous studies, older adults show a reduction in the HbR concentration as well as an increase in the HbO_2_ concentration in dual-task walking, which indicates an increase in the prefrontal area activation (Hawkins et al., 2018, Hoang et al., 2022, Izzetoglu and Holtzer, 2020, Salzman et al., 2021b). Nevertheless, the outcomes of this study’s data synthesis are not comparable to those of previous studies. Despite HbR has been utilized as a measure of brain activity, fNIRS typically imposes limitations on the detection of HbR spectra (Leff et al., 2011). Due to the low amplitude of HbR spectra, distinguishing between single-task and dual-task is more difficult (Leff et al., 2011). In addition, the low amplitude is more likely to cause interference with tissue surrounding the head, including the skull, and frontal sinus, and cerebrospinal fluid (Leff et al., 2011). Consequently, the absence of a significant variation in HbR concentration during dual-task walking may be attributable to the aforementioned constraints.

### 4.3 Clinical implications

Utilizing brain hemodynamics, particularly under dual-task paradigms, may provide an early detection approach for screening risk of falling in older adults. Early detection of alterations in cortical hemodynamics may also provide a significant chance to design an appropriate intervention, thereby minimizing the risk of falling among older adults.

### 4.4 Study limitations

Several limitations of the present study must be taken into account when interpreting the findings. First, this meta-analysis study did not restrict the types of cognitive tasks that were included, in order to analyze the differential effects of complexity of dual-tasking. Future studies and reviews should elucidate the influence of these factors on cortical activity. Second, the cognitive tasks used in some studies involved speaking, for instance, counting backwards and verbal fluency requiring muscles which are located nearby the PFC (Zimeo Morais et al., 2018). Such muscle activities and different facial expressions may affect fNIRS signal quality (Balardin et al., 2017). Third, the studies in our systemic review were conducted using healthy older adults. Therefore, our conclusions cannot be generalized to patients with cognitive impairment (i.e., dementia, Alzheimer’s disease) or motor deficits (i.e., post-stroke, Parkinson’s disease). Therefore, future studies that examine the associations between falling in older adults and dual-tasking should be conducted in different patient groups.

## 5. Conclusions

Older adults exhibit an increase in oxyhemoglobin concentration in the PFC when performing the dual-task paradigms, suggesting an increase in cortical activity. The increase in cortical activity might explain why older adults are more likely to fall while walking, walk slowly, or reduce physical activity. Although cognitive tasks (e.g., alternate letters, serial subtractions, and general conversation) are not particularly difficult, older persons spend considerable effort to accomplish tasks, particularly those requiring inhibitory control. Therefore, we propose that cognitive tasks associated with inhibitory control tasks are more likely to influence cortical activity during dual-task walking.

## Data Availability

The data from this study are available on request from the corresponding author.

## Institutional review board statement

This is a meta-analysis study. No ethical approval is required.

## Informed consent statement

This is a meta-analysis study. The informed consent is not applicable.

## Data availability statement

The data from this study are available on request from the corresponding author.

## Supplementary materials Search strategy

PubMed (n = 336)

((“elderly”[Title/Abstract]) OR (“older”[Title/Abstract])) AND ((“cognit*”[Title/Abstract]) OR (“dual*”[Title/Abstract])) AND ((“fNIRS”[Title/Abstract]) OR (“hemodynamic”[Title/Abstract]))

Web of science (n = 391)

(AB=(“elderly”) OR AB=(older)) AND (AB=(“cognit*”) OR AB=(“dual*”)) AND (AB=(“fNIRS”) OR AB=(“hemodynamic”))

Scopus (n = 693)

(TITLE-ABS-KEY (“elderly”) OR TITLE-ABS-KEY (“older”)) AND (TITLE-ABS-KEY (“cognit*”) OR TITLE-ABS-KEY (“dual*”)) AND (TITLE-ABS-KEY (“fNIRS”) OR TITLE-ABS-KEY (“hemodynamic”)) AND (LIMIT-TO (DOCTYPE, “ar”))

## References

Abbott, R. D., et al. 2004. Walking and dementia in physically capable elderly men. Jama, 292, 1447–53. https://doi.org/10.1001/jama.292.12.1447.

Atkinson, H. H., et al. 2007. Cognitive function, gait speed decline, and comorbidities: the health, aging and body composition study. J Gerontol A Biol Sci Med Sci, 62, 844–50. https://doi.org/10.1093/gerona/62.8.844.

Balardin, J. B., et al. 2017. Imaging Brain Function with Functional Near-Infrared Spectroscopy in Unconstrained Environments. Front Hum Neurosci, 11. https://doi.org/10.3389/fnhum.2017.00258.

Beauchet, O., et al. 2008. Recurrent falls and dual task-related decrease in walking speed: is there a relationship? J Am Geriatr Soc, 56, 1265–9. https://doi.org/10.1111/j.1532-5415.2008.01766.x.

Beauchet, O., et al. 2009. Stops walking when talking: a predictor of falls in older adults? Eur J Neurol, 16, 786–95. https://doi.org/10.1111/j.1468-1331.2009.02612.x.

Beurskens, R., et al. 2014. Age-related changes in prefrontal activity during walking in dual-task situations: a fNIRS study. Int J Psychophysiol, 92, 122–8. https://doi.org/10.1016/j.ijpsycho.2014.03.005.

Bootsma-Van Der Wiel, A., et al. 2003. Walking and talking as predictors of falls in the general population: the Leiden 85-Plus Study. J Am Geriatr Soc, 51, 1466–71. https://doi.org/10.1046/j.1532-5415.2003.51468.x.

Cabeza, R., Nyberg, L. 2000. Imaging cognition II: An empirical review of 275 PET and fMRI studies. J Cogn Neurosci, 12, 1–47. https://doi.org/10.1162/08989290051137585.

Callan, D. E., et al. 2014. Multisensory and modality specific processing of visual speech in different regions of the premotor cortex. Front Psychol, 5, 389. https://doi.org/10.3389/fpsyg.2014.00389.

Chaparro, G., et al. 2017. Frontal brain activation changes due to dual-tasking under partial body weight support conditions in older adults with multiple sclerosis. J Neuroeng Rehabil, 14, 65. https://doi.org/10.1186/s12984-017-0280-8.

Chen, M., et al. 2017. Neural correlates of obstacle negotiation in older adults: An fNIRS study. Gait Posture, 58, 130–135. https://doi.org/10.1016/j.gaitpost.2017.07.043.

Chen, Y., et al. 2022. Increased cortical activation and enhanced functional connectivity in the prefrontal cortex ensure dynamic postural balance during dual-task obstacle negotiation in the older adults: A fNIRS study. Brain Cogn, 163, 105904. https://doi.org/10.1016/j.bandc.2022.105904.

Corp, D. T., et al. 2018. Reduced motor cortex inhibition and a ‘cognitive-first’ prioritisation strategy for older adults during dual-tasking. Exp Gerontol, 113, 95–105. https://doi.org/10.1016/j.exger.2018.09.018.

De Hoon, E. W., et al. 2003. Quantitative assessment of the stops walking while talking test in the elderly. Arch Phys Med Rehabil, 84, 838–42. https://doi.org/10.1016/s0003-9993(02)04951-1.

Diamond, A. 2013. Executive functions. Annu Rev Psychol, 64, 135–68. https://doi.org/10.1146/annurev-psych-113011-143750.

Dietrich, A., Audiffren, M. 2011. The reticular-activating hypofrontality (RAH) model of acute exercise. Neurosci Biobehav Rev, 35, 1305–25. https://doi.org/10.1016/j.neubiorev.2011.02.001.

Fraser, S. A., et al. 2017. Does combined physical and cognitive training improve dual-task balance and gait outcomes in sedentary older adults? Front Hum Neurosci, 10. https://doi.org/10.3389/fnhum.2016.00688.

Gabrieli, J. D., et al. 1998. The role of left prefrontal cortex in language and memory. Proc Natl Acad Sci U S A, 95, 906–13. https://doi.org/10.1073/pnas.95.3.906.

George, C. J., et al. 2019. The effect of polypharmacy on prefrontal cortex activation during single and dual task walking in community dwelling older adults. Pharmacol Res, 139, 113–119. https://doi.org/10.1016/j.phrs.2018.11.007.

Grady, C. L. 2000. Functional brain imaging and age-related changes in cognition. Biol Psychol, 54, 259–281. https://doi.org/10.1016/S0301-0511(00)00059-4.

Hall, C. D., et al. 2010. Efficacy of gaze stability exercises in older adults with dizziness. J Neurol Phys Ther, 34, 64–9. https://doi.org/10.1097/NPT.0b013e3181dde6d8.

Hassan, S. A., et al. 2020. Changes in Oxyhemoglobin Concentration in the Prefrontal Cortex during Cognitive-Motor Dual Tasks in People with Chronic Obstructive Pulmonary Disease. Copd, 17, 289–296. https://doi.org/10.1080/15412555.2020.1767561.

Hausdorff, J. M. 2005. Gait variability: methods, modeling and meaning. J Neuroeng Rehabil, 2, 19. https://doi.org/10.1186/1743-0003-2-19.

Hawkins, K. A., et al. 2018. Prefrontal over-activation during walking in people with mobility deficits: Interpretation and functional implications. Hum Mov Sci, 59, 46–55. https://doi.org/10.1016/j.humov.2018.03.010.

Hernandez, A. R., et al. 2020. A Cross-species Model of Dual-Task Walking in Young and Older Humans and Rats. Front Aging Neurosci, 12, 276. https://doi.org/10.3389/fnagi.2020.00276.

Hernandez, M. E., et al. 2016. Brain activation changes during locomotion in middle-aged to older adults with multiple sclerosis. J Neurol Sci, 370, 277–283. https://doi.org/10.1016/j.jns.2016.10.002.

Hoang, I., et al. 2022. Increased prefrontal activity during usual walking in aging. Int J Psychophysiol, 174, 9–16. https://doi.org/10.1016/j.ijpsycho.2022.01.011.

Holtzer, R., et al. 2018. The effect of diabetes on prefrontal cortex activation patterns during active walking in older adults. Brain Cogn, 125, 14–22. https://doi.org/10.1016/j.bandc.2018.03.002.

Holtzer, R., Izzetoglu, M. 2020. Mild Cognitive Impairments Attenuate Prefrontal Cortex Activations during Walking in Older Adults. Brain Sci, 10. https://doi.org/10.3390/brainsci10070415.

Holtzer, R., et al. 2019a. Distinct fNIRS-Derived HbO_2_ Trajectories During the Course and Over Repeated Walking Trials Under Single- and Dual-Task Conditions: Implications for Within Session Learning and Prefrontal Cortex Efficiency in Older Adults. J Gerontol A Biol Sci Med Sci, 74, 1076–1083. https://doi.org/10.1093/gerona/gly181.

Holtzer, R., et al. 2019b. The effect of fear of falling on prefrontal cortex activation and efficiency during walking in older adults. Geroscience, 41, 89–100. https://doi.org/10.1007/s11357-019-00056-4.

Holtzer, R., et al. 2011. fNIRS study of walking and walking while talking in young and old individuals. J Gerontol A Biol Sci Med Sci, 66, 879–87. https://doi.org/10.1093/gerona/glr068.

Holtzer, R., et al. 2020. Intraindividual variability in neural activity in the prefrontal cortex during active walking in older adults. Psychol Aging, 35, 1201–1214. https://doi.org/10.1037/pag0000583.

Holtzer, R., et al. 2022. Cognitive Reserve Moderates the Efficiency of Prefrontal Cortex Activation Patterns of Gait in Older Adults. J Gerontol A Biol Sci Med Sci, 77, 1836–1844. https://doi.org/10.1093/gerona/glab288.

Holtzer, R., et al. 2016. Neurological Gait Abnormalities Moderate the Functional Brain Signature of the Posture First Hypothesis. Brain Topogr, 29, 334–43. https://doi.org/10.1007/s10548-015-0465-z.

Izzetoglu, M., Holtzer, R. 2020. Effects of Processing Methods on fNIRS Signals Assessed During Active Walking Tasks in Older Adults. IEEE Trans Neural Syst Rehabil Eng, 28, 699–709. https://doi.org/10.1109/tnsre.2020.2970407.

Kannus, P., et al. 2005. Aging and Degeneration of Tendons. In: Maffulli, N., Renström, P. & Leadbetter, W. B. (eds.) Tendon Injuries: Basic Science and Clinical Medicine. London: Springer London.

Karim, H. T., et al. 2013. Neuroimaging to detect cortical projection of vestibular response to caloric stimulation in young and older adults using functional near-infrared spectroscopy (fNIRS). Neuroimage, 76, 1–10. https://doi.org/10.1016/j.neuroimage.2013.02.061.

Leff, D. R., et al. 2011. Assessment of the cerebral cortex during motor task behaviours in adults: a systematic review of functional near infrared spectroscopy (fNIRS) studies. Neuroimage, 54, 2922–36. https://doi.org/10.1016/j.neuroimage.2010.10.058.

Li, K. Z., et al. 2001. Walking while memorizing: age-related differences in compensatory behavior. Psychol Sci, 12, 230–7. https://doi.org/10.1111/1467-9280.00341.

Lindenberger, U., et al. 2000. Memorizing while walking: increase in dual-task costs from young adulthood to old age. Psychol Aging, 15, 417–36. https://doi.org/10.1037//0882-7974.15.3.417.

Lundin-Olsson, L., et al. 1997. “Stops walking when talking” as a predictor of falls in elderly people. Lancet, 349, 617. https://doi.org/10.1016/s0140-6736(97)24009-2.

Luo, D., et al. 2018. Optimally estimating the sample mean from the sample size, median, midrange, and/or mid-quartile range. Stat Methods Med Res, 27, 1785–1805. https://doi.org/10.1177/0962280216669183.

Maidan, I., et al. 2016. The Role of the Frontal Lobe in Complex Walking Among Patients With Parkinson’s Disease and Healthy Older Adults: An fNIRS Study. Neurorehabil Neural Repair, 30, 963–971. https://doi.org/10.1177/1545968316650426.

Marusic, U., et al. 2019. Aging effects on prefrontal cortex oxygenation in a posture-cognition dual-task: an fNIRS pilot study. Eur Rev Aging Phys Act, 16, 2. https://doi.org/10.1186/s11556-018-0209-7.

Maruya, K., et al. 2021. Brain Activity in the Prefrontal Cortex during Cognitive Tasks and Dual Tasks in Community-Dwelling Elderly People with Pre-Frailty: A Pilot Study for Early Detection of Cognitive Decline. Healthcare (Basel), 9. https://doi.org/10.3390/healthcare9101250.

Mihara, M., et al. 2008. Role of the prefrontal cortex in human balance control. Neuroimage, 43, 329–36. https://doi.org/10.1016/j.neuroimage.2008.07.029.

Mirelman, A., et al. 2017. Effects of aging on prefrontal brain activation during challenging walking conditions. Brain Cogn, 115, 41–46. https://doi.org/10.1016/j.bandc.2017.04.002.

Page, M. J., et al. 2021. The PRISMA 2020 statement: an updated guideline for reporting systematic reviews. BMJ, 372, n71. https://doi.org/10.1136/bmj.n71.

Pakray, H., et al. 2021. The Effects of Perceived Pain in the Past Month on Prefrontal Cortex Activation Patterns Assessed During Cognitive and Motor Performances in Older Adults. Pain Med, 22, 303–314. https://doi.org/10.1093/pm/pnaa404.

Pinti, P., et al. 2020. The present and future use of functional near-infrared spectroscopy (fNIRS) for cognitive neuroscience. Ann N Y Acad Sci, 1464, 5–29. https://doi.org/10.1111/nyas.13948.

Quaresima, V., Ferrari, M. 2019. Functional Near-Infrared Spectroscopy (fNIRS) for Assessing Cerebral Cortex Function During Human Behavior in Natural/Social Situations: A Concise Review. Organ Res Methods, 22, 46–68. https://doi.org/10.1177/1094428116658959.

Ross, D., et al. 2021. Prefrontal cortex activation during dual-task walking in older adults is moderated by thickness of several cortical regions. Geroscience, 43, 1959–1974. https://doi.org/10.1007/s11357-021-00379-1.

Rosso, A. L., et al. 2017. Neuroimaging of an attention demanding dual-task during dynamic postural control. Gait Posture, 57, 193–198. https://doi.org/10.1016/j.gaitpost.2017.06.013.

Sabharwal, S., et al. 2015. Heterogeneity of the definition of elderly age in current orthopaedic research. Springerplus, 4, 516. https://doi.org/10.1186/s40064-015-1307-x.

Salzman, T., et al. 2021a. Prefrontal Cortex Involvement during Dual-Task Stair Climbing in Healthy Older Adults: An fNIRS Study. Brain Sci, 11. https://doi.org/10.3390/brainsci11010071.

Salzman, T., et al. 2021b. Hemodynamic and behavioral changes in older adults during cognitively demanding dual tasks. Brain Behav, 11, e02021. https://doi.org/10.1002/brb3.2021.

Shi, J., et al. 2020. Optimally estimating the sample standard deviation from the five-number summary. Res Synth Methods, 11, 641–654. https://doi.org/10.1002/jrsm.1429.

St George, R. J., et al. 2021. Functional Near-infrared Spectroscopy Reveals the Compensatory Potential of Pre-frontal Cortical Activity for Standing Balance in Young and Older Adults. Neuroscience, 452, 208–218. https://doi.org/10.1016/j.neuroscience.2020.10.027.

Sterne, J. A., et al. 2016. ROBINS-I: a tool for assessing risk of bias in non-randomised studies of interventions. Bmj, 355, i4919. https://doi.org/10.1136/bmj.i4919.

Stuart, S., et al. 2019. Monitoring multiple cortical regions during walking in young and older adults: Dual-task response and comparison challenges. Int J Psychophysiol, 135, 63–72. https://doi.org/10.1016/j.ijpsycho.2018.11.006.

Talamonti, D., et al. 2022. Prefrontal hyperactivation during dual-task walking related to apathy symptoms in older individuals. PLoS One, 17, e0266553. https://doi.org/10.1371/journal.pone.0266553.

Talamonti, D., et al. 2021. The Benefits of Physical Activity in Individuals with Cardiovascular Risk Factors: A Longitudinal Investigation Using fNIRS and Dual-Task Walking. J Clin Med, 10. https://doi.org/10.3390/jcm10040579.

Teo, W. P., et al. 2021. Altered prefrontal cortex responses in older adults with subjective memory complaints and dementia during dual-task gait: An fNIRS study. Eur J Neurosci, 53, 1324–1333. https://doi.org/10.1111/ejn.14989.

Verghese, J., et al. 2002. Validity of divided attention tasks in predicting falls in older individuals: a preliminary study. J Am Geriatr Soc, 50, 1572–6. https://doi.org/10.1046/j.1532-5415.2002.50415.x.

Verghese, J., et al. 2007. Quantitative gait dysfunction and risk of cognitive decline and dementia. J Neurol Neurosurg Psychiatry, 78, 929–35. https://doi.org/10.1136/jnnp.2006.106914.

Wagshul, M. E., et al. 2019. Multi-modal neuroimaging of dual-task walking: Structural MRI and fNIRS analysis reveals prefrontal grey matter volume moderation of brain activation in older adults. Neuroimage, 189, 745–754. https://doi.org/10.1016/j.neuroimage.2019.01.045.

Weuve, J., et al. 2004. Physical activity, including walking, and cognitive function in older women. Jama, 292, 1454–61. https://doi.org/10.1001/jama.292.12.1454.

Wu, T., et al. 2004. How self-initiated memorized movements become automatic: a functional MRI study. J Neurophysiol, 91, 1690–8. https://doi.org/10.1152/jn.01052.2003.

Young, W. R., et al. 2016. Examining links between anxiety, reinvestment and walking when talking by older adults during adaptive gait. Exp Brain Res, 234, 161–72. https://doi.org/10.1007/s00221-015-4445-z.

Zimeo Morais, G. A., et al. 2018. fNIRS Optodes’ Location Decider (fOLD): a toolbox for probe arrangement guided by brain regions-of-interest. Scientific Reports, 8, 3341. https://doi.org/10.1038/s41598-018-21716-z.

